# Assessment of gender and geographical bias in the editorial decision-making process of biomedical journals: A Case-Control study

**DOI:** 10.1101/2024.03.15.24304220

**Authors:** Angèle Gayet-Ageron, Khaoula Ben Messaoud, Mark Richards, Cyril Jaksic, Julien Gobeill, Jeevanthi Liyanapathirana, Luc Mottin, Nona Naderi, Patrick Ruch, Zoé Mariot, Alexandra Calmy, Julia Friedman, Leonard Leibovici, Sara Schroter

**Author notes:** equal contribution (Angèle Gayet-Ageron, Khaoula Ben Messaoud). Correspondence to: Angèle Gayet-Ageron. (https://orcid.org/0000-0002-6164-9693), (https://orcid.org/0000-0002-8053-4672), (https://orcid.org/0000-0001-8689-5711), (https://orcid.org/0000-0001-5835-3480), (https://orcid.org/0000-0001-9809-7741), (https://orcid.org/0000-0002-9614-9293), (https://orcid.org/0000-0002-1272-7640), (https://orcid.org/0000-0002-3374-2962), (https://orcid.org/0009-0000-9238-888X), (https://orcid.org/0000-0002-1137-6826), (https://orcid.org/0000-0002-9907-344X), (https://orcid.org/0000-0002-8791-8564).

## Abstract

**Objectives:** To assess whether the gender (primary) and geographical affiliation (post hoc) of the first and/or last authors of manuscripts is associated with publication decisions after controlling for known confounders.

**Design:** Case-control (1:1) study.

**Setting:** Two large general medical journals and 20 specialist journals.

**Participants:** Original peer reviewed research manuscripts submitted between January 1, 2012, and December 31, 2019.

**Main outcomes measures and predictor:** Manuscripts accepted (cases) and rejected (controls) were compared between women and men first authors (main predictor), and between women and men last authors (secondary predictor).

**Results:** Of 7,000 included manuscripts 6,724 (96.1%) first and 6,768 (96.7%) last authors’ gender were identified; 3,056 (43.7%) and 2,214 (32.7%) were women, respectively. The proportion of women first and last authors were respectively 46.7% (n=1,571) and 32.3% (n=1,093) among cases and 44.2% (n=1,485) and 33.1% (n=1,121) among controls. In univariate analysis, being a woman first author increased the likelihood of acceptance for publication (odds ratio 1.11; 95% confidence interval 1.00 to 1.22). After adjustment for study attributes, then post-hoc variables, the association between the first author’s gender and acceptance for publication became non-significant 1.04 (0.94 to 1.16). The likelihood of acceptance for publication was significantly lower for first authors affiliated to Asia 0.58 (0.48 to 0.70) compared to Europe, and for first author affiliated to upper middle income 0.61 (0.47 to 0.78) and lower middle and low-income 0.65 (0.45 to 0.93) compared to high income countries. Compared to papers where both first and last authors were from the same country, acceptance for publication was significantly higher when both authors were affiliated to different countries from the same geographical and income groups 1.39 (1.09 to 1.77), to different countries, different geographical but same income groups 1.45 (1.14 to 1.84), or to different income groups 1.59 (1.20 to 2.11). The study attributes (design, and funding) were also independently associated with acceptance for publication.

**Conclusions:** The absence of gender inequalities during the editorial decision-making process is reassuring. However, the underrepresentation of first authors affiliated to Asia and low-income countries in manuscripts accepted for publication indicates poor representation of global scientists’ opinion and supports growing demands for improving diversity in biomedical research.

**Summary boxes:** *What is already known on this topic?:*

- Published studies have revealed gender inequalities in academia, attainment of leadership positions, successful grant funding and in biomedical research production (including reported research contributions, attainment of key authorship positions in submitted and published manuscripts, membership of editorial boards, and participation in peer review).
- Geographical inequalities have been reported in biomedical research production, including membership of editorial boards, participation in peer review, and attainment of key authorship positions in submitted and published articles.
- Diversity in science is associated with better research quality and improved representation of global scientists’ opinions.

*What this study adds?:*

- Gender inequality toward the first author of original research manuscripts disappeared after adjusting for other important factors related to authorship (first authors’ geographical affiliation and income group), and the study attributes (funding and medical specialty).
- First authors affiliated to Asia compared to Europe and from low-income compared to high-income countries had a lower chance of being accepted for publication.
- Geographical and income diverse teams represented by the first and the last authors’ affiliations had a greater chance of being accepted for publication.

## Introduction

The editorial decision-making process should be impartial and based on the quality of manuscripts, compliance with the journal’s requirements and relevance for its readership; the study results and authors’ characteristics should not be part of the decision to publish. Yet, biases in editorial decision-making have been reported.(1–3) Studies have shown that editorial decision-making is influenced by the methodology used in the reported study (such as the study design, the sample size and the statistical methods used), (4,5) the strength of the findings, (6); the source of funding, (4) positive findings, (7) and whether the corresponding author is affiliated to the same country as the editor of the journal. (4) Editorial decisions can further be influenced by the perceived high citation potential of the research in a highly competitive editorial market (8,9) and potential reprint sales revenue (10).

It is not known whether first and/or last authors’ gender is associated with the chance of getting published. Authors’ gender has been shown to be associated with the reported types of contribution to research projects (11) and position on the authorship byline of submitted manuscripts (12). Moreover, men first authors are more likely to present research findings positively in the titles and abstracts of their articles compared to women first authors (13) and this could potentially influence editorial decisions in favour of men first authors. Although some studies concluded to the absence of implicit gender biases after finding no difference on acceptance between anonymised vs non-anonymised peer review processes, they were assessed in a single journal specialised in ecology.(14)

The objectives of the ATHENA case-control study were to assess in a large sample of biomedical journals whether there is an association between the gender of the first (main objective) and/or last author (secondary objective), and acceptance for publication independently of known factors related to the methods and funding of the research described in the manuscript.

Recently, the geographical location of researchers’ affiliation has been identified as being associated with knowledge dissemination, including authorship positions, publication rates, citation counts and invitation to peer review (15–17). Geographical bias plays a role in the assessment of the quality of research articles and helps explain the under-representation of low– and middle-income country authors in published articles and subsequently in the citations generated by this research.(15) The lack of diversity among editors and peer reviewers in terms of gender, geographical affiliation, income group, race, and ethnicity could also contribute to the paucity of diversity in research publications.(16,17) Because these studies were published after the writing of our original protocol but prior to our data analyses, we added a new secondary objective to test whether geographical affiliation and country’s income of authors was associated with acceptance for publication (Figure 1), as this outcome had not been specifically studied yet.

**Figure 1.**
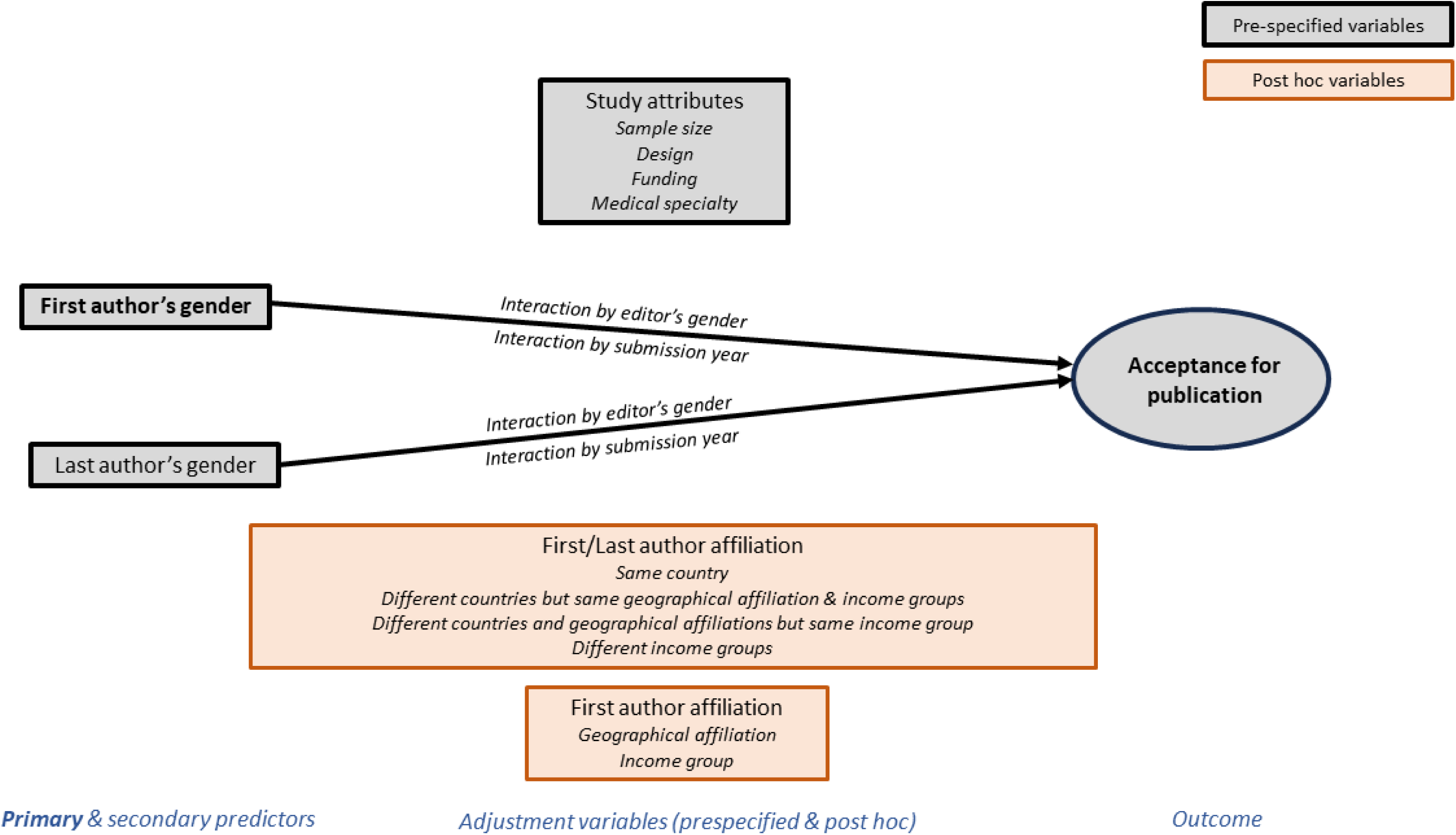
Conceptual framework of the ATHENA project including primary/secondary predictors, pre-specified and post hoc adjustment variables to model acceptance for publication.

## Methods

### Study design and settings

The ATHENA study was a case-control study nested in a prospective cohort study of all original research manuscripts submitted for publication in 22 biomedical journals (21 from the BMJ Publishing Group, and one from Elsevier – [Clinical Microbiology & Infection, CMI], Supplementary Table 1 and Figure 1).

### Participants and data source

We included all original research articles and systematic reviews submitted to 22 participating journals between January 1, 2012, and December 31, 2019, and sent out for peer review, and which had received a final editorial decision by 28 February 2022. Among eligible articles, we randomly sampled our cases and controls from the BMJ Publishing Group database and the CMI database. We extracted data from the manuscript submission systems for BMJ Publishing Group journals and we received data directly from the CMI database for this specific journal. Data related to the characteristics of the original research reported in these manuscripts were automatically extracted using machine learning from the submitted PDFs by investigators from the Swiss Institute for Bioinformatics (PR, NN, LM, JL, JG).

### Case and control definition

Cases were manuscripts accepted for publication with at least one completed peer review report. Controls were manuscripts rejected for publication with at least one completed peer review report. Accepted and rejected articles (1:1) were randomly selected at the journal level.

### Exposure factors explored and other variables

For each submitted manuscript we collected details about the journal (impact factor (<5, 5-10, >10) and type of peer review process (open vs. anonymised); manuscript (manuscript ID, title, abstract, original submission date, first decision date, number of co-authors, editorial decision); handling editor (first name, last name, country of affiliation); authors (authorship position (where several co-authors held the first or last authorship position we used the first and last declared author respectively), corresponding author status, salutation, first name, middle name, last name, country of affiliation). We classified the countries of affiliation into six geographical affiliation groups (Europe, Africa, Asia, South America, North America or Oceania) and then further categorised this by country wealth using the four levels defined by the World Bank Atlas method according to 2021’s gross national income per capita (low income; lower middle income; upper middle income; and high income).(18) For CMI manuscripts we did not receive data on authors’ and editors’ country of affiliation, middle names, nor salutation.

#### Gender determination (main and secondary predictors)

Editors’ gender was provided by BMJ Publishing Group and was estimated for CMI using Gender API based on editor’s first name alone. We used a four-step sequential process to determine authors’ gender. Firstly, we used the first name and country of affiliation (except for CMI) in Gender API (https://gender-api.com/en) website. Gender API provides gender determination with an accuracy probability from 50% to 100% (under 50% an unknown status is attributed). Secondly, for undetermined gender we used middle names and the country of affiliation in Gender API (except for CMI). Thirdly, we used the online service genderize.io (http://genderize.io) to determine gender based on first and middle names (except for CMI) which also considers gender as unknown if accuracy is under 50%. Fourthly (except for CMI), we used authors’ salutation and attributed with 100% accuracy man to “Mr” or “M” and woman to “Miss,” “Mrs,” and “Ms.”. Where determined gender had an accuracy below 80%, we retained the gender determined by salutation and attributed an accuracy of 100%. Finally, we determined gender at three levels of accuracy: ≥60%, ≥70% and ≥80% for all authors and reviewers. We used accuracy ≥60% for all analyses. We cross-tabulated the gender of first and last authors and created the variable “gender diversity between first and last author”.

### Adjustment variables

We estimated the percentage of women authors among all authors on the byline with a determined gender at three levels of accuracy: ≥60%, ≥70% and ≥80%, and again used accuracy ≥60% for all analyses. We created a 4-category variable: no women authors,0 to 49%, 50% to 99%, and 100% women authors.

We created a “Diversity of authors’ affiliations” variable using the country, geographical affiliation group, and country income group of the first and last authors (post hoc variables). We identified four groups: 1) first and last authors affiliated to the same country; 2) first and last authors affiliated to different countries within the same geographical affiliation and income groups; 3) first and last authors had same geographical affiliation and income groups; 4) first and last author had different income groups.

#### Study attributes

To identify medical specialty, funding type, study design and sample size of the study reported in each submitted manuscript, we used machine learning procedures detailed in Supplementary Materials 2-5 and Supplementary Table 2

### Study sample size estimation

We chose an odds ratio (OR) of 0.85 to indicate a presence of gender bias towards women for the association between first author’s gender and acceptance for publication. We assumed the proportion of women first authors to be 40% among cases (44% among controls) using OR=0.85. Considering a type-1 error at 5% (two-sided) with a study power of 90% and a correlation coefficient for exposure between matched controls and cases at 0.03 (to consider some clustering at the journal level), we approximated that 2,800 cases and 2,800 controls were needed. Anticipating 20% missing values or exclusions we increased our sample size to 3500 cases/controls (1:1). We took a random selection of cases and controls by journal, considering 159 journal-years across the study period and 20 cases/controls per journal-year corresponding to a maximum of 160 cases/controls per journal.

### Statistical analysis

We present continuous variables as means (standard deviations, SD) or medians (25^th^ and 75^th^ percentiles), categorical variables as frequencies and relative frequencies by case/control status. We describe data for CMI journal separately in Supplementary Table 3 as we did not have all the data for primary analysis. For a randomly selected set of 100 manuscripts, we evaluated the level of agreement between the studies’ attributes as automatically extracted by machine learning procedures– leveraging the SIB Literature Services (SIBiLS) (18)– compared to a manual extraction performed by consensus between three independent raters (AGA, KBM, ZM). We calculated unweighted kappa coefficients (ᴋ) for the variables medical specialty, and design; weighted kappa coefficient (ᴋw) for the variables funding and sample size. Primary and secondary analyses were conducted for BMJ Publishing Group manuscripts only. For manuscripts with at least 2 co-authors, we performed unconditional logistic regression models on complete cases with case/control status as the dependent variable. The main predictor was the first author’s gender, and the last author’s gender was a secondary predictor. Initially, the gender of the corresponding author was a secondary predictor, however, in 81% of cases (n=1,282), they also held the position of first or last author, so we did not test it. First authors’ gender, prespecified variables (last author’s gender, study attributes) then post hoc variables (geographical affiliation group and income group of first authors, and diversity of authors’ affiliation) were introduced in the main model. The conceptual framework for the selection of variables is shown in Figure 1. First authors’ gender was introduced first in the model, followed by the other prespecified variables (last author’s gender, study attributes), then the post hoc variables (geographical affiliation group, income group of first authors, and diversity of authors’ affiliation). We prespecified four interaction terms between (1) gender of the editor and first author; (2) gender of the editor and last author; (3) submission year and gender of the first author; (4) submission year and gender of the last author. We report unadjusted and adjusted odds ratios, 95% confidence intervals (95% CI), and p-values from univariate and multivariable models. We performed three additional sensitivity analyses using levels of accuracy of ≥70%, and ≥80% for gender determination, and a multiple imputation model for missing gender of the first and last authors. Missing gender of the first and last authors were considered at random, and 20 imputations were applied; adjustment of the imputation model was done for geographical affiliation group of the first and last authors, and for the journal. For categorical predictors with more than two modalities, a global p-value was computed. We interpreted global p-values as significant if p<0.05. As a post-hoc analysis, we calculated the likelihood ratio of the null hypothesis of no gender bias over the alternative hypothesis of presence of gender bias for the association between first author’s gender and acceptance for publication (19). We used Stata intercooled (STATA Corp., College Station, Texas, USA) 17.0 for analyses and R software (version 4.1.2) for data management.

### Patient and public involvement

We partnered and co-authored with the chair of BMJ’s LGBTQ+ network (MR). MR contributed to data collection, gender determination method, manuscript revision, and final approval.

## Results

We included 7,000 manuscripts from BMJ Publishing Group with all pre-specified variables available for analysis (Figure 2, Table 1). We found a moderate level of agreement between raters and automatic algorithm extraction on medical specialty identification (ᴋ=0.58), funding type (ᴋw=0.54), study design (ᴋ=0.33) and study sample size (ᴋw=0.84).

**Figure 2.**
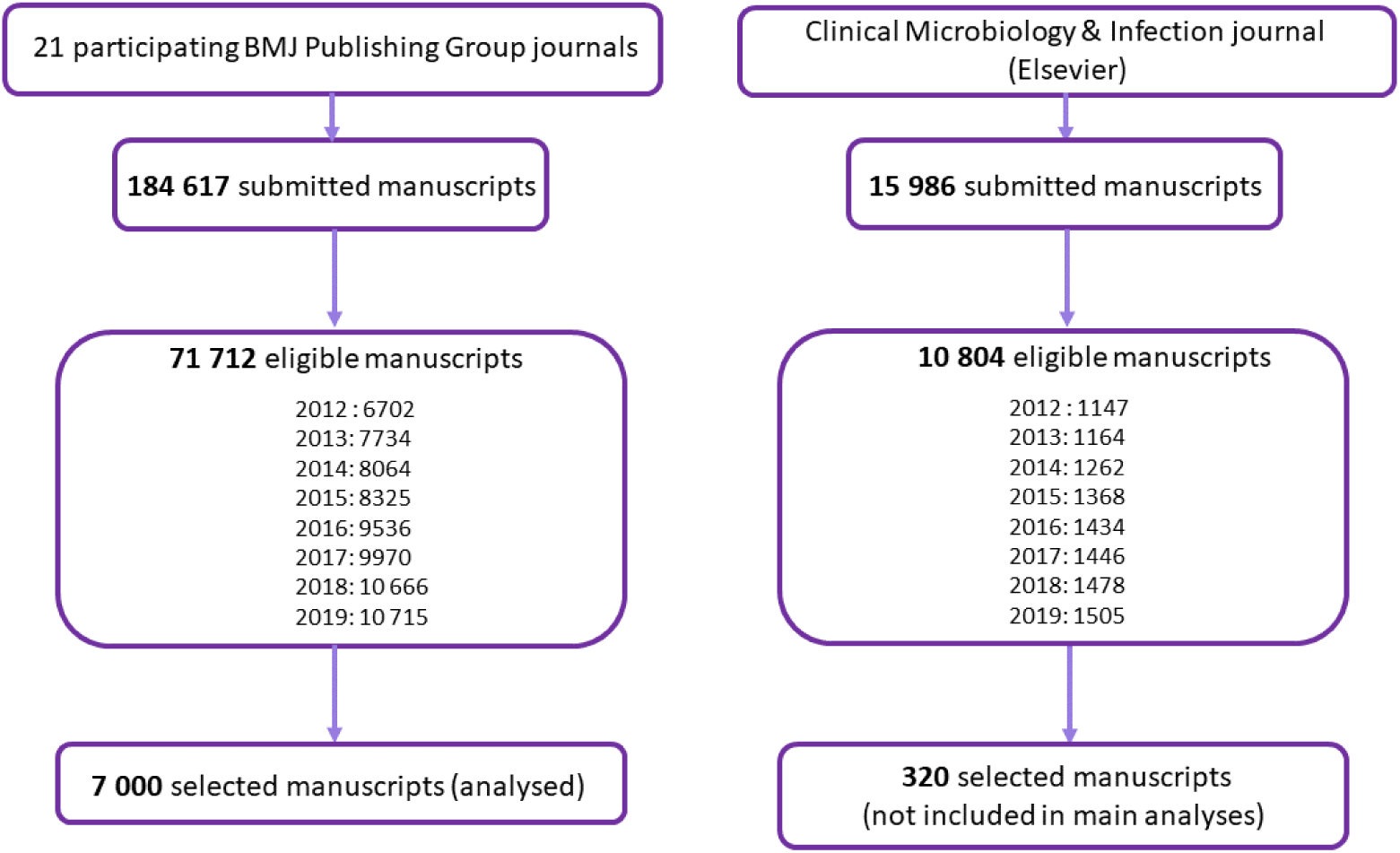
Description of all manuscripts submitted, eligible and selected for ATHENA study between January 1, 2012 and December 31, 2019 by publisher.

**Table 1.**
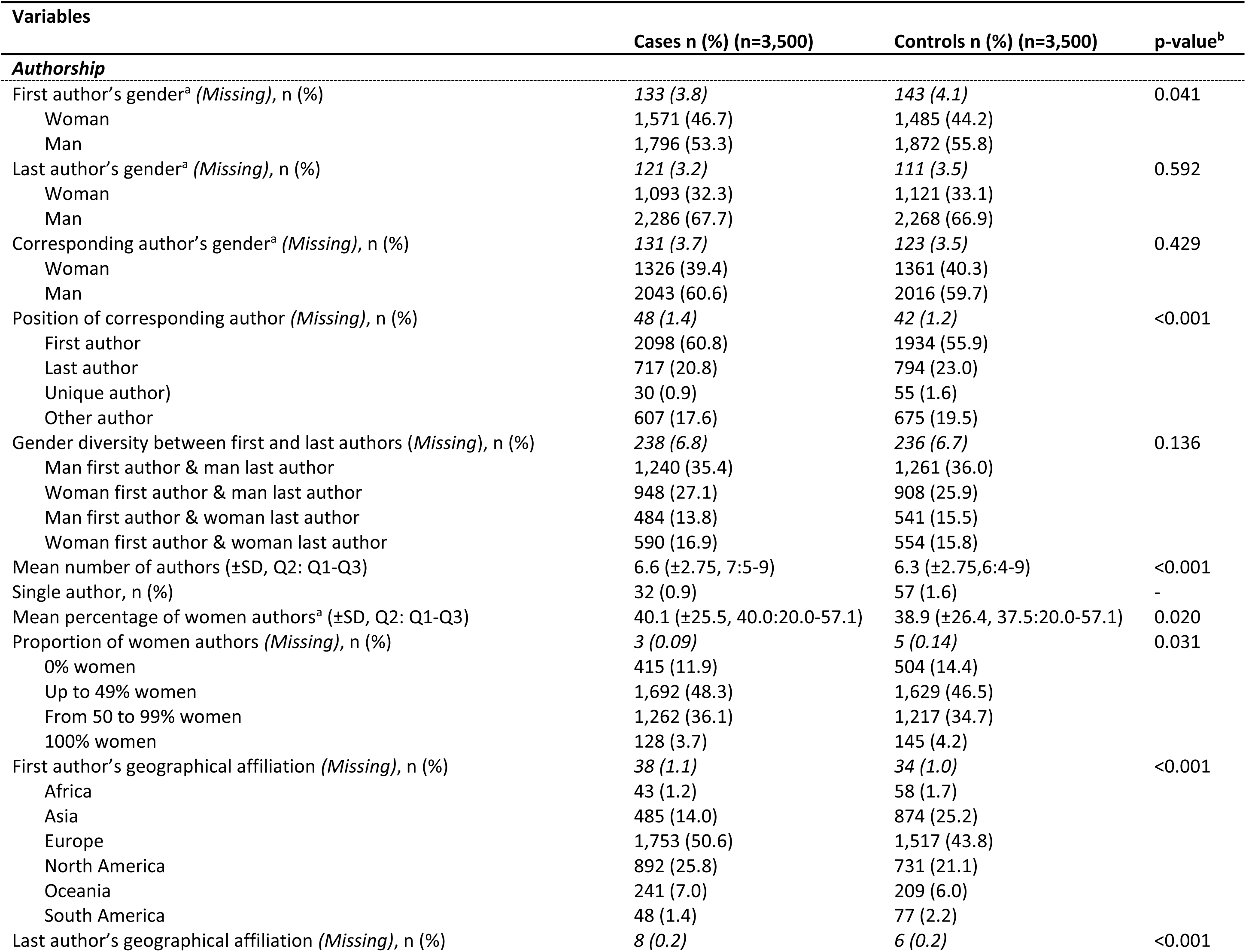
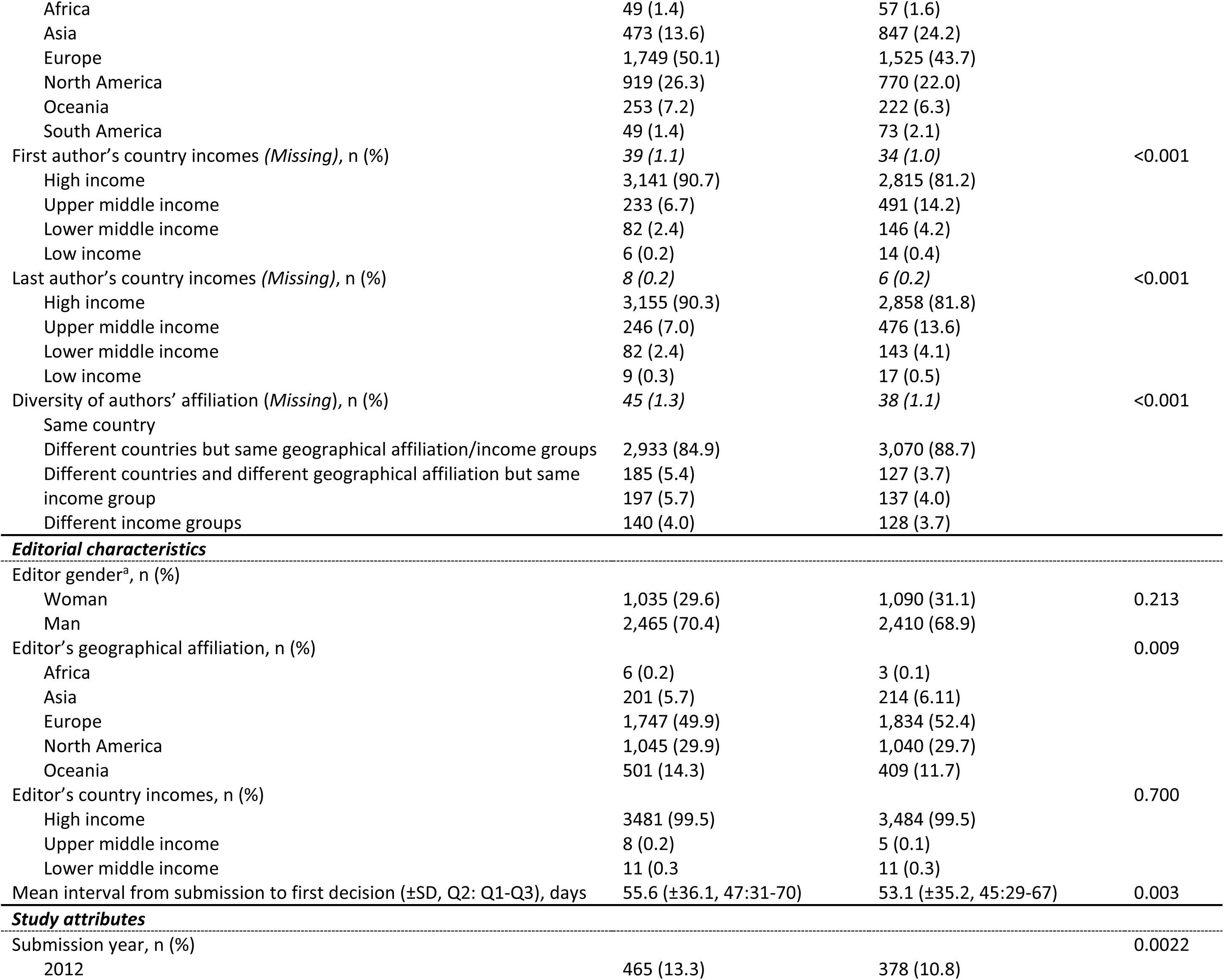
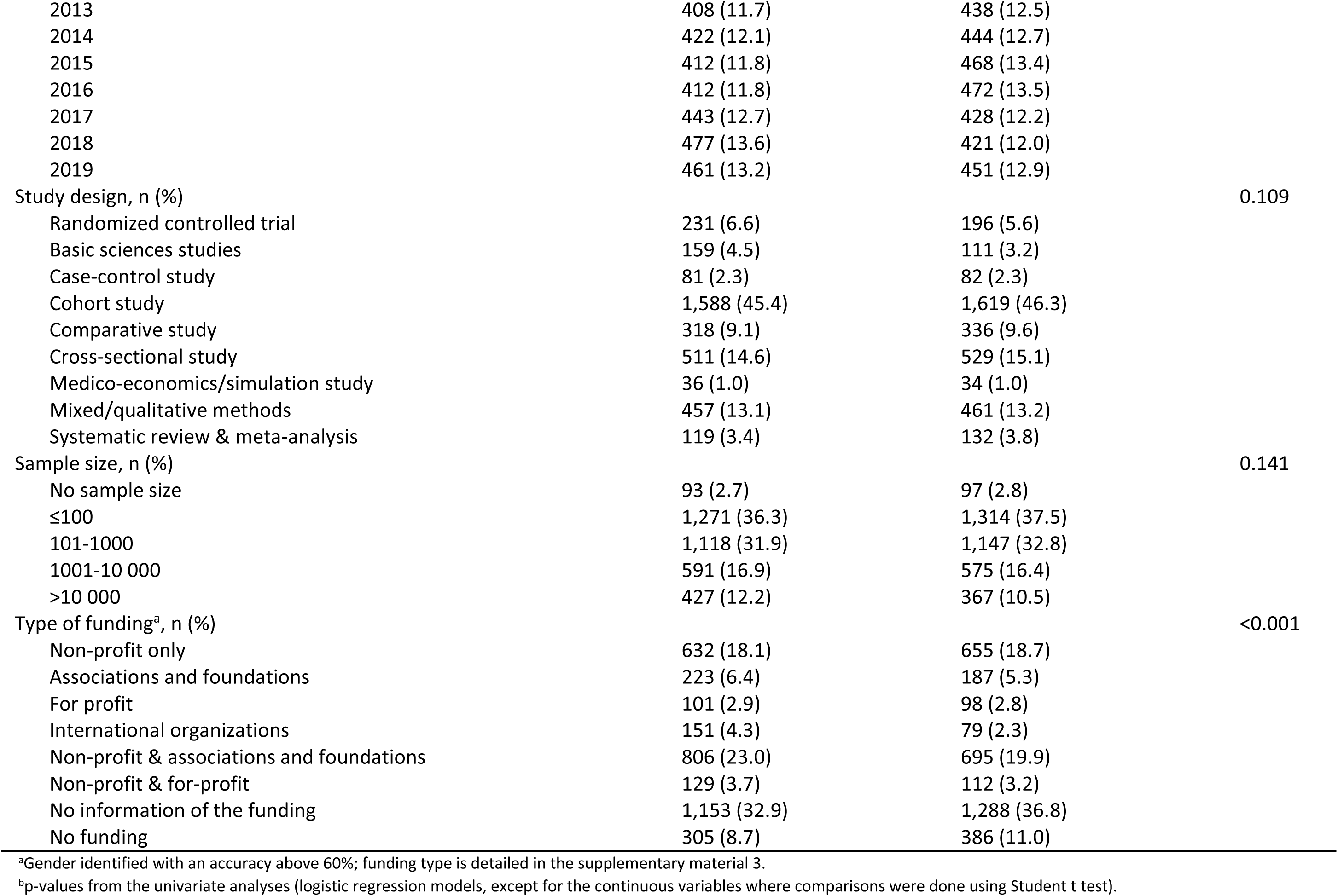
Proportion of editorial, authorship, and study attributes for 21 BMJ Publishing Group journals by case and control status.

### Proportion of editorial, authorship and manuscript characteristics by accepted and rejected status

The distribution of editor’s geographical affiliation was significantly different between accepted and rejected manuscripts, and the mean duration from submission to first decision was on average 2.5 days longer for accepted than rejected (Table 1). The proportion of women first authors was 2.5 points higher, and the overall percentage of women co-authors 1.2% higher in accepted than in rejected, both differences were statistically significant between accepted and rejected. Accepted manuscripts had on average +0.3 co-authors compared to rejected, and more rejected than accepted had only one author. There were significantly more first or last authors with European, North American, or Oceanian affiliations and they were more often affiliated with a high-income country among accepted than rejected. The proportion of manuscripts with different geographical affiliation groups or income groups between first and last authors was significantly higher for accepted compared to rejected. Regarding the manuscript attributes, the proportion of studies stating no funding or where funding was not declared were significantly lower among accepted than rejected. The proportion of accepted manuscripts was significantly higher in 2012 compared to later years.

### Association between gender of the authors and acceptance for publication (primary objective)

Table 2 shows univariate and multivariable analyses of the main analysis. The association between gender of the first author and acceptance for publication became non-significant once funding, and medical specialty were introduced in the model; it stayed non-significant after adjustment for the post hoc variables (geographical affiliation group and income group of first authors and diversity of authors’ affiliations). Gender of the last author was non-significantly associated with acceptance for publication in the univariate and multivariable analyses. The interaction between the gender of the editor and first author (p=0.158), and between the gender of the editor and last author (p=0.160) were both non-significant. The interactions between the year of submission and respectively the gender of first (p=0.637) and last authors (p=0.662) were non-significant. We found similar findings with the sensitivity analyses (Supplementary Table 4). Acceptance for publication was also independently associated with study design (higher acceptance for basic science studies compared with clinical trials), study sample size (higher for studies with >10,000 participants compared to ≤100 or no sample size), and with type of funding (higher for studies funded by international organisations, non-profit & associations & foundations, and associations & foundations compared to studies declaring no funding) (Table 2, and Figure 2). The likelihood ratio of no gender bias (null hypothesis) over existence of gender bias (alternative hypothesis) was 909, providing decisively strong evidence in favour of the null hypothesis.

**Table 2:** Acceptance for publication in a random sample of 7000 accepted/rejected manuscripts submitted to 21 BMJ Publishing Group journals between January 1, 2012 and December 31, 2019 with at least two coauthors on the byline. Univariate and multivariable models ^a,b^.

| Independent variables | Univariate analyses |  |  | Multivariable analysis<sup>a,b</sup> |  |  |
| --- | --- | --- | --- | --- | --- | --- |
|  | OR | 95%CI | p-value | OR | 95%CI | p-value |
| **Author / Editor characteristics** |  |  |  |  |  |  |
| First author's gender<sup>c</sup> (ref= Man) |  |  | 0.041 |  |  | 0.413 |
| Woman | 1.11 | (1.00 to 1.22) |  | 1.04 | (0.94 to 1.16) |  |
| Last author's gender<sup>c</sup> (ref= Man) |  |  | 0.592 |  |  | 0.652 |
| Woman | 0.97 | (0.88 to 1.08) |  | 0.98 | (0.88 to 1.09) |  |
| Gender diversity between first and last authors (ref=men first & last) |  |  | 0.136 |  |  | - |
| Woman first author & man last author | 1.06 | (0.94-1.20) | 0.347 | - | - | - |
| Man first author & woman last author | 0.91 | (0.78-1.05) | 0.198 | - | - | - |
| Woman first author & woman last author | 1.09 | (0.95-1.26) | 0.227 | - | - | - |
| Proportion of women co-authors on byline<sup>c</sup> (ref= only men) |  |  | 0.031 | - | - | - |
| Up to 49% women | 1.24 | (1.07 to 1.44) | 0.005 | - | - | - |
| From 50 to 99% women | 1.24 | (1.06 to 1.45) | 0.007 | - | - | - |
| 100% women | 1.15 | (0.86 to 1.52) | 0.346 |  |  |  |
| First author's geographical affiliation<sup>e</sup> (ref=Europe) |  |  | <0.001 |  |  | <0.001 |
| Africa | 0.65 | (0.44 to 0.97) | 0.036 | 0.91 | (0.54 to 1.52) | 0.716 |
| Asia | 0.48 | (0.42 to 0.55) | <0.001 | 0.58 | (0.48 to 0.70) | <0.001 |
| North America | 1.06 | (0.94 to 1.20) | 0.312 | 1.07 | (0.94 to 1.22) | 0.296 |
| Oceania | 1.00 | (0.82 to 1.22) | 0.990 | 1.05 | (0.86 to 1.30) | 0.623 |
| South America | 0.55 | (0.38 to 0.79) | 0.001 | 0.78 | (0.50 to 1.23) | 0.283 |
| First author's country of affiliation income (ref=high income) |  |  | <0.001 |  |  | <0.001 |
| Upper middle income | 0.42 | (0.36 to 0.50) | <0.001 | 0.61 | (0.47 to 0.78) | <0.001 |
| Lower middle and low income | 0.51 | (0.39 to 0.67) | <0.001 | 0.65 | (0.45 to 0.93) | 0.019 |
| Last author's geographical affiliation (ref=Europe) |  |  |  |  |  | - |
| Africa | 0.76 | (0.52 to 1.12) | 0.169 | - | - | - |
| Asia | 0.49 | (0.43 to 0.56) | <0.001 | - | - | - |
| North America | 1.05 | (0.93 to 1.18) | 0.439 | - | - | - |
| Oceania | 1.00 | (0.82 to 1.21) | 0.994 | - | - | - |
| South America | 0.59 | (0.41 to 0.86) | 0.005 | - | - | - |
| Last author's country of affiliation income (ref=high income) |  |  | <0.001 |  |  | - |
| Upper middle income | 0.46 | (0.39 to 0.55) | <0.001 | - | - | - |
| Lower middle and low income | 0.54 | (0.41 to 0.70) | <0.001 | - | - | - |
| Geographical/income diversity between first/last authors (ref=same country) |  |  | <0.001 |  |  | <0.001 |
| Different countries, but same geographical affiliation/income group | 1.51 | (1.20 to 1.91) | <0.001 | 1.39 | (1.09 to 1.77) | 0.008 |
| Different countries and geographical affiliation but same income group | 1.49 | (1.19 to 1.87) | <0.001 | 1.45 | (1.14 to 1.84) | 0.002 |
| Different income groups | 1.14 | (0.89 to 1.45) | 0.308 | 1.59 | (1.20 to 2.11) | 0.001 |
| Editor's gender<sup>c</sup> (ref= Man) |  |  | 0.213 |  |  |  |
| Woman | 0.94 | (0.85 to 1.04) |  | - | - | - |
| Editor's geographical affiliation (ref=Europe) |  |  | 0.009 |  |  |  |
| Africa | 2.09 | (0.52 to 8.38) | 0.297 | - | - | - |
| Asia | 1.00 | (0.82 to 1.24) | 0.959 | - | - | - |
| North America | 1.06 | (0.95 to 1.18) | 0.306 | - | - | - |
| Oceania | 1.30 | (1.12 to 1.51) | <0.001 | - | - | - |
| Editor's country of affiliation income (ref=high income) |  |  | 0.700 |  |  | - |
| Upper middle income | 1.59 | (0.52 to 4.86) | 0.417 | - | - | - |
| Lower middle income | 0.90 | (0.38 to 2.13) | 0.816 | - | - | - |
| **Study attributes** |  |  |  |  |  |  |
| Submission year (ref=2012)) |  |  | 0.0022 |  |  | - |
| 2013 | 0.74 | (0.61 to 0.90) | 0.003 | - | - | - |
| 2014 | 0.77 | (0.63 to 0.93) | 0.007 | - | - | - |
| 2015 | 0.71 | (0.59 to 0.86) | <0.001 | - | - | - |
| 2016 | 0.70 | (0.69 to 1.01) | <0.001 | - | - | - |
| 2017 | 0.83 | (0.75 to 1.10) | 0.060 | - | - | - |
| 2018 | 0.91 | (0.68 to 0.99) | 0.341 | - | - | - |
| 2019 | 0.82 | (0.68 to 0.99) | 0.044 | - | - | - |
| Study design (ref= Randomized controlled trial) |  |  | 0.109 |  |  | 0.045 |
| Basic sciences studies | 1.21 | (0.89 to 1.65) | 0.224 | 1.43 | (1.02 to 2.01) | 0.040 |
| Case-control study | 0.82 | (0.57 to 1.19) | 0.295 | 0.87 | (0.59 to 1.28) | 0.483 |
| Cohort study | 0.83 | (0.68 to 1.02) | 0.076 | 0.85 | (0.68 to 1.06) | 0.149 |
| Comparative study | 0.80 | (0.63 to 1.03) | 0.084 | 0.90 | (0.69 to 1.18) | 0.448 |
| Cross-sectional study | 0.83 | (0.66 to 1.04) | 0.112 | 0.83 | (0.65 to 1.06) | 0.137 |
| Medico-economics/simulation study | 0.95 | (0.57 to 1.59) | 0.841 | 0.86 | (0.50 to 1.48) | 0.580 |
| Mixed/qualitative methods | 0.84 | (0.67 to 1.06) | 0.134 | 0.83 | (0.65 to 1.07) | 0.148 |
| Systematic review & meta-analysis | 0.75 | (0.55 to 1.03) | 0.077 | 0.83 | (0.59 to 1.16) | 0.276 |
| Sample size (ref= ≤100 or no sample size) |  |  | 0.141 |  |  | 0.064 |
| 101-1000 | 1.00 | (0.90 to 1.12) | 0.959 | 1.07 | (0.95 to 1.21) | 0.275 |
| 1001-10000 | 1.06 | (0.92 to 1.22) | 0.408 | 1.11 | (0.95 to 1.29) | 0.198 |
| >10000 | 1.19 | (1.02 to 1.40) | 0.029 | 1.28 | (1.07 to 1.53) | 0.008 |
| Type of funding (ref. No funding) |  |  | <0.001 |  |  | <0.001 |
| No information of funding | 1.11 | (0.93 to 1.31) | 0.250 | 1.07 | (0.89 to 1.28) | 0.489 |
| Non-profit | 1.18 | (0.98 to 1.43) | 0.078 | 1.19 | (0.98 to 1.46) | 0.085 |
| Associations and foundations | 1.47 | (1.15 to 1.88) | 0.002 | 1.47 | (1.14 to 1.92) | 0.004 |
| For-profit | 1.25 | (0.91 to 1.72) | 0.165 | 1.13 | (0.81 to 1.58) | 0.484 |
| International organizations | 2.41 | (1.76 to 3.29) | <0.001 | 2.30 | (1.64 to 3.23) | <0.001 |
| Non-profit & associations & foundations | 1.43 | (1.19 to 1.71) | <0.001 | 1.31 | (1.08 to 1.60) | 0.007 |
| Non-profit & for-profit | 1.40 | (1.04 to 1.88) | 0.025 | 1.30 | (0.95 to 1.78) | 0.102 |

<sup>a</sup>Model performed for manuscripts with more than one author (n=6,428 observations). <sup>b</sup>Model adjusted for the speciality of the research topic (p=0.327). <sup>c</sup>Gender determined with accuracy ≥60%.

### Association between geographical affiliation of the authors and acceptance for publication (post hoc objective)

Acceptance for publication was significantly associated with geographical affiliation group of the first author: compared to Europe, the adjusted odds ratio was significantly lower for first authors affiliated to Asia (Table 2). The likelihood of acceptance was significantly lower when first author was affiliated to upper-middle or lower-middle and low-income groups. We found similar results when using the affiliation group of the last author, except that the odds for publication acceptance was not statistically different for the comparison between Africa and Europe (data not shown). When high income group was the reference, the odds ratio was lower for first authors affiliated to upper-middle, lower middle, and low-income group or countries. Compared to when first and last authors were affiliated to the same countries, the odds ratio was higher than 1 when both were affiliated to different countries but the same geographical and income groups, when both were affiliated to different countries and geographical affiliations but same income group or when both had different income groups (Table 2).

## Discussion

Manuscripts with a woman first author were not associated with lower manuscript acceptance as hypothesised. Nor was the gender of the last author associated with acceptance for publication. Post-hoc calculation of the likelihood ratio provided decisive evidence supporting the absence of gender bias in publication acceptance. In the additional analyses, the probability of being accepted for publication was lower when the first author was affiliated to an Asian country than a European country, and when first author was affiliated with an upper-middle or lower-middle/low-income compared to high income country. Nevertheless, the geographical diversity among the first and last authors’ affiliation was associated with a higher probability of acceptance. Study design and type of funding were also independently associated with acceptance for publication.

### Comparison with other studies

Previous studies (4, 20, 21, 22) also failed to show an association between the gender of key authorship positions and acceptance for publication. Those studies were mainly based on a single medical specialty and used different research methods none of which adjusted for the author and study attributes (23). Similar to our findings, Burns et al. demonstrated that papers with a first author affiliated to Asia, Africa, or South America had a lower chance of acceptance for publication than those with authors affiliated to North America. (22) The impact of the diversity of authors’ affiliations on manuscript outcomes, such as acceptance for publication, has been little studied in science. However, Campbell et al explored submissions and publications across American Geophysical Union journals from 2012-2018 in terms of national affiliation, gender, career stage of individual authors and race/ethnicity for US-based authors (24). Acceptance was +2.8% higher for cross-cultural collaborations compared to international teams, and +4.5% higher for mixed-gender compared to single-gender teams. In line with our findings, they found no significant difference in acceptance rates between manuscripts with a woman or man first author. Although they used a more intersectional method for defining diversity, they too found diverse teams to have higher acceptance rates than non-diverse teams.

### Study strengths and limitations

The inclusion of submissions from 21 journals over 8 years, including two large general medical journals, and journals in a range of biomedical specialties attracting submissions from around the world provided a rich and diversified sample. While inclusion of journals from BMJ Publishing Group alone, which has a commitment to improving equality and diversity (25), limits the generalisability of the findings to other journals, we sampled manuscripts submitted from 2012 to 2019. Back then, equality, diversity and inclusion were less of a focus for publishers than in recent years (25,26), particularly since the Royal Society of Chemistry launched the Joint Commitment for Action on Inclusion and Diversity in Publishing in 2020 in response to the Black Lives Matter movement.(27) Although our study initially planned to include more biomedical journals from other publishers, we only managed to include one other journal which could not provide the same set of data as the BMJ Publishing Group so we presented this data separately.

To limit information bias, we chose a case-control design and randomly selected cases and controls from eligible manuscripts using a computerised procedure. In the absence of self-identified data from authors, we determined gender as binary and by doing so may have misrepresented people with diverse gender identities. However, we applied a four step sequential procedure already implemented in our two previous works (12,16) and used the threshold accuracy of gender determination above 60% for our primary analysis. Our three sensitivity analyses using higher threshold accuracies for gender determination and multiple imputation to replace missing gender data were in line with our original results. Unlike previous studies (23), we adjusted for author and study attributes which are important confounders. However, we acknowledge that the quality of a study and its manuscript is a major factor influencing publication acceptance and could be associated with many of our predictors, making it an important confounding variable. However, in the absence of a rigorous tool we could not robustly assess manuscript quality for such a large set of manuscripts. We used an algorithm trained with machine learning to automatically extract study attributes but the agreement between raters and the automatic algorithm extraction of variables was only moderate. Discrepancies between manual and automatic extractions were randomly distributed and should not have biased the estimated association between study attributes and the outcome. Defining diversity is complex as it incorporates various intersectional dimensions. We explored a definition of author team diversity by combining information from two covariates (geographical area and income group of the first and last authors’ country of affiliation) and by adjusting for their gender. This definition is limited as it is not based on the affiliation of all co-authors and only included the first institutional affiliation for an author if there were multiple so teams may have been more diverse than what we captured.

### Policy implications

The fact that we did not identify gender inequalities during the editorial decision-making process is reassuring but gender bias in research production still deserves further attention, as gender inequalities have been demonstrated for submitted and published articles particularly during the early covid-19 pandemic.(12) Our results need to be confirmed in a randomised trial where the influence of known and unknown confounding factors can be accounted for. The limited geographical representation among first authors in accepted manuscripts heightens the potential for an inadequate reflection of the opinions of global scientists. More crucially, it may also result in a lack of representation of patients and populations requiring prevention and care.(15,16) The determinants and implications of lower acceptance of manuscripts from Asia, and from low-income countries need to be further explored. Our finding of a positive effect on publication acceptance of geographically and gender diverse teams has been demonstrated by others.(28–31) Subjective and hidden factors, such as implicit cognitive biases related to geographical affiliations, might also drive perceptions of study quality and the judgements of peer reviewers and subsequently the final editorial decision, however, they are more difficult to investigate.(32) Anonymising peer review has been proposed to reduce bias against authors but a meta-analysis has shown it does not affect the quality of peer reviewers’ reports. (33) Moreover, open and published peer review aims to restore trust in the integrity and fairness of the review process through transparency to readers has also been shown not to affect the quality of review.(34) In addition, double anonymising the peer review process has not been shown to improve geographic diversity among authors(35), but triple-anonymised peer review (where even the handling editor of a manuscript cannot see the identity of the authors) has provided some evidence of reducing editorial bias.(36) Educating faculty members about implicit biases and strategies for overcoming them has provided positive effects on implicit biases surrounding women and leadership in academic medicine(37) and should be extended to geographical implicit bias. As gatekeepers of publication decisions, editors have a crucial role to play in improving global diversity in research through the acknowledgement of their own potential implicit biases by monitoring sensitive diversity indicators for their peer reviewers and the authors of manuscripts they choose to reject or peer review. They can also promote the systemic detection of implicit biases in reviewers’ comments, associate editors’ decisions and the content of the authors’ research itself as recently advocated by *Nature* (39). Linguistic bias is another barrier for non-native English-speaking researchers, especially from low-or middle-income countries, and can lower the chance of acceptance for publication.(34) Other possible solutions to facilitate the publication of high-quality research in regional languages include the decentralisation of editorial boards, by including for example Spanish or Chinese board members, publishing articles in regional languages, or decentralising the indexing of journals by creating independent citation databases at regional level (for e.g. the African Citation Index).(40)Provision of free or low-cost translation and support services promoted by high-income universities or other public academic institutions and designed for non-native English speaking authors is thus desirable.(41) Some journals are starting to experiment with AI-powered scientific writing assistants (42) and uptake and beneficial effects of using these should be monitored.

### Conclusions

Absence of gender bias in the final editorial decision is reassuring. Research needs diverse teams (43) and equitable partnerships with low resource countries (44) yet inclusivity in biomedical research is currently insufficient. Funders, academic institutions, researchers, biomedical journal editors and publishers are key to achieving this goal.

## Data Availability

Data described in the manuscript, code book, and analytic code will be archived and preserved on Yareta, the research data repository for Geneva higher education institutions. Data will not be made publicly available, but relevant anonymised data are available on reasonable request from the corresponding author. The study was conducted under a confidentiality agreement between BMJ Publishing Group and the medical school of Geneva University represented by the Department of Health and Community Medicine and the Swiss Institute for Bioinformatics and the HES-SO/HEG Geneva.

## Acknowledgements

We thank the editors of the participating journals from the BMJ Publishing Group for giving us access to the data. We thank Professor J. Peter Donnelly, editor-in-chief and Colin Drummond for having evaluated the potential participation of the Journal of Antimicrobial Chemotherapy in the ATHENA case-control study. We thank Prof. Martin Richard Tramèr for having provided a dataset for pilot study from the European Journal of Anaesthesiology, as well as M. Marco De Ambrogi from the Lancet Infectious Diseases. We thank Prof. Thomas Perneger for providing useful advice on the presentation of the results.

## Ethical approval

Ethical approval was received from the Institutional Review Board of the Geneva Canton on April 16, 2018 for the ATHENA project (N° 2019-00540).

## Data sharing

Data described in the manuscript, code book, and analytic code will be archived and preserved on Yareta, the research data repository for Geneva’s higher education institutions. Data will not be made publicly available, but relevant anonymised data are available on reasonable request from the corresponding author. The study was conducted under a confidentiality agreement between BMJ Publishing Group and the medical school of Geneva University represented by the Department of Health and Community Medicine and the Swiss Institute for Bioinformatics and the HES-SO – HEG Genève.

## Contributors

The ATHENA study was funded by the SNSF. AGA, LL, SS, and AC were involved in the study concept and design of the ATHENA study. AGA (BMJ Publishing Group journals) and LL (CMI journal) were responsible for the recruitment of the participating journals. SS (BMJ Publishing Group) and JF (CMI) were responsible for data collection; PR, JL, NN, JG, LM (Swiss Institute for Bioinformatics & HES-SO – HEG Genève) and CJ were responsible for data extraction. KBM was responsible for updating the study method under AGA supervision. MR and KBM were responsible for defining the methods for the gender determination. KBM determined the gender. KBM was responsible for data management. AGA, KBM, and ZM were responsible for validation of the automated data extraction. AGA was responsible for obtaining research funding. AGA and KBM were involved in data analysis and interpretation. AGA generated the figures; AGA and KBM the tables. AGA wrote the initial draft manuscript and managed the versions. KBM provided the draft of the introduction and the method sections. All authors were involved in critical revision of multiple drafts and approved the final version. AGA is the guarantor. The corresponding author attests that all listed authors meet authorship criteria and that no others meeting the criteria have been omitted.

