## Supplemental Materials for "Assessment of gender and geographical bias in the editorial decision-making process of biomedical journals: A Case-Control study"

### Table of contents


**Section 1: Supplementary details on methods**

**Supplementary Table 1**: Description of participating biomedical journals.

**Supplemental material 2:** Strategy for identifying medical specialties from the participating journals.

**Supplementary material 3.** Strategy for identifying research funding.

**Supplementary material 4.** Strategy for extracting the study design.

**Supplementary Table 2.** MeSH headings and number of publications found in SIBiLS for each design label.

**Supplementary material 5.** Strategy for extracting the sample size.

**Section 2: Supplementary descriptive analysis**

**Supplementary Table 3:** Description of cases and controls by editorial, authorship, and study characteristics for the CMI journal (n=320).

**Section 3: Supplementary multivariable analyses**

**Supplementary Table 4:** Acceptance for publication in a random sample of 7,000 accepted/rejected manuscripts submitted to 21 BMJ Publishing Group journals between January 1, 2012 and December 31, 2019 with at least two coauthors on the byline. Multivariable analyses from three sensitivity analyses.

**Supplementary Table 1.** Description of participating biomedical journals.

| Journals name | Impact factor (2019 JCR) | Impact factor class | Type of peer review process | Generalist or specialist journal |
|---|---|---|---|---|
| **BMJ Publishing Group** | | | | |
| *The BMJ* | 30.31 | >10 | Open | Generalist |
| *BMJ Global Health* | 4.28 | <5 | Single anonymised | Specialist |
| *BMJ Open* | 2.5 | <5 | Open | Generalist |
| *Gut* | 19.82 | >10 | Single anonymised | Specialist |
| *Heart* | 5.21 | 5-10 | Single anonymised | Specialist |
| *Journal of Neurology, Neurosurgery, and Psychiatry* | 8.26 | 5-10 | Single anonymised | Specialist |
| *Occupational and Environmental Medicine* | 3.82 | <5 | Single anonymised | Specialist |
| *Postgraduate Medical Journal* | 1.91 | <5 | Single anonymised | Specialist |
| *Sexually Transmitted Infections* | 3.4 | <5 | Single anonymised | Specialist |
| *Thorax* | 10.84 | >10 | Single anonymised | Specialist |
| *Tobacco Control* | 6.95 | 5-10 | Single anonymised | Specialist |
| *BMJ Quality & Safety* | 7.035 | 5-10 | Triple anonymised | Specialist |
| *British Journal of Ophthalmology* | 4.638 | <5 | Single anonymised | Specialist |
| *Journal of Medical Genetics* | 6.318 | 5-10 | Single anonymised | Specialist |
| *Journal of Clinical Pathology* | 3.411 | <5 | Single anonymised | Specialist |
| *Journal of NeuroInterventional Surgery* | 5.836 | 5-10 | Double anonymised | Specialist |
| *Archives of Disease in Childhood* | 3.801 | <5 | Single anonymised | Specialist |
| *BMJ Paediatrics Open* | NA[a] | <5 | Open | Specialist |
| *Annals of the Rheumatic Diseases* | 19.103 | >10 | Single anonymised | Specialist |
| *Journal of Epidemiology & Community Health* | 3.71 | <5 | Single anonymised | Specialist |
| *British Journal of Sports Medicine* | 13.8 | >10 | Single anonymised | Specialist |
| **Oxford University Press Inc.** | | | | |
| *Clinical Microbiology & Infection* | 2.942 | <5 | Single anonymised | Specialist |

[a]Impact factor not available (NA) in 2019. We attribute impact factor class <5 to *BMJ Paediatrics Open*.

**Supplementary material 2.** Strategy for identifying medical specialties from the participating journals.

Three sequential strategies were used to obtain medical speciality. First, we assigned a speciality based on journal name for the following specialty journals: *BMJ Paediatrics Open* (Paediatrics & Neonatology), *Heart* (Cardiology), *Journal of Medical Genetics* (Genetics), *Journal of Neurology, Neurosurgery and Psychiatry* (Neurology), *British Journal of Ophthalmology* (Ophthalmology), *British Journal of Sports Medicine* (Sports medicine). For the remaining journals, we explored keywords, primary subject headings, titles, and abstracts of the selected manuscripts to guess the assigned MeSH terms and therefore the most likely related specialty. Finally, we trained an automatic multi-class classifier using a Support Vector Machine (SVM) using MEDLINE data. Given an input text, the system outputs a list of specialties ranked by confidence score.

We used a total of 24 medical speciality categories, as follows: Cardiology, Dermatology & Venereology, Emergency & Perioperative Medicine, Endocrinology, Environmental Medicine, Epidemiology, Gastroenterology, and Genetics, Global Health, Gynaecology & Obstetrics, Immunology, Infectious Disease Medicine, Internal Medicine, Neurology, Oncology, Ophthalmology, Paediatrics & Neonatology, Psychiatry, Psychology & Addictology, Pulmonary Medicine, Radiology & Connected Medicine, Rheumatology, Sports Medicine, Surgery.

**Supplementary material 3.** Strategy for identifying research funding.

Identifying the categories of funding sources necessitated crafting a pipeline that combines information extraction with the Crossref database, enabling the identification of funding types, including those from public entities, private companies, and foundations. The final system is a multiclass multilabel classifier, which given an article attempt to identify a passage containing the funding information to output the type of the funding source.

The first step involved extracting the text containing funding information from the data set. We first converted the PDF files of the data set to a structured XML/TEI ile using the GROBID (https://github.com/kermitt2/grobid) PDF parsing software. The resulting files in XML/TEI format are then automatically screened to find text patterns, important keywords, and section titles likely to indicate the source of funding. Based on these findings, we determined which passages of the text should then be used to extract the funding data.

In parallel, we accessed the most recent update (12/2022) of the Crossref funder registry (https://www.crossref.org/services/funder-registry/), which contains 31,182 entries of funding entities. Each entry includes a DOI number, the preferred label or name for the funding entity, and optionally some alternative labels, including synonyms and acronyms. This expanded our list to 91,083 funding entities along with their metadata. Thanks to the DOI, we queried the CrossrefAPI (https://www.doi.org/) to obtain the funding type for each entity. Indeed, all entities belong to one of following ten categories: Associations and Societies, For-profit companies, International Organizations, Libraries and data archiving organizations, Local Government, National Government, Other non-profit organizations, Research Institutes and centers, Trusts/Charities/Foundations, and Universities.

We combined both steps mentioned above together, to obtain the necessary funding information. In some articles, the funding source was either not available or not identified by our pipeline, therefore an 11$^{th}$ category was created: “No funding information”.

We further clustered the funding body types into eight categories, as follows: 1) Non-profit organizations (including Universities, National Government, Local Government, Research Institutes and centers, Libraries and data archiving organizations or Other non-profit organizations), 2) Associations and societies (private and public) & Trusts/Charities/Foundations (both public and private), 3) For-profit companies (including pharmaceutical industry), 4) International Organizations, 5) Non-profit organizations & Associations/Societies & Trusts/Charities/Foundations, 6) Non-profit organizations & For-profit companies, 7) No funding (when there is no funding entity found), and 8) No information of the funding (when no information related to funding was found).

**Supplementary material 4.** Strategy for extracting the study design.

Experimental designs refer to the methodological frameworks that researchers use to plan, conduct, and analyse experiments. To automatically extract the study design from documents, we built a 12-class classifier. We used the MeSH terminology to normalize these designs and to train a machine learning model using a large data set of MEDLINE articles. The MeSH headings were used to extract 142'566 MEDLINE records in September 2022, from SIBiLS [1] (see Supplemental Table 2 below).

**Supplemental Table 2.** MeSH headings and number of publications found in SIBiLS for each design label.

| Experimental designs | MeSH headings | MeSH IDs | Nb of publications |
|---|---|---|---|
| cross-sectional study | Cross-Sectional Studies | D003430 | 11,147 |
| qualitative study | Qualitative Research | D036301 | 2,511 |
| animal study | Animals ;<br>Animal Experimentation | D000818 ;<br>D032761 | 19,710 |
| systematic review & meta-analysis | Systematic Reviews as Topic ;<br>Systematic Review ;<br>Meta-Analysis as Topic ;<br>Meta-Analysis | D000078202 ;<br>D000078182 ;<br>D015201 ;<br>D017418 | 8,030 |
| cohort study | Cohort Studies | D015331 | 8,458 |
| randomized controlled trial | Randomized Controlled Trials as Topic ;<br>Randomized Controlled Trial | D016032 ;<br>D016449 | 15,775 |
| comparative study | Comparative Study | D003160 | 21,007 |
| epidemiological method | Epidemiologic Methods | D004812 | 1,914 |
| case-control study | Case-Control Studies | D016022 | 7,199 |
| simulation analysis | Computer Simulation | D003198 | 377 |
| medico-economics study | Cost-Benefit Analysis | D003362 | 2,132 |
| case reports | Case Reports | D002363 | 44,306 |
| mixed methods study | - | - | - |

We then used this collection to train a GradientBoost model, which was empirically selected for its capacity to provide reliable results that cope with relatively small unbalanced datasets [2], adjusting the training parameters of the model to overcome this possible learning bias. For each document, we ask our classifier to provide the top two labels, with their associated confidence scores. On the test set, our model achieved an overall accuracy of 78% for the primary label prediction, which further improved to 90% when considering the second prediction as well.

This result prompted us to apply the following rule when extracting study designs from manuscripts: If the accuracy of the primary label prediction is <0.35, and the difference between accuracies of the two labels is <0.05, we retain the design with the second highest accuracy. Else, we select the label with the highest estimated accuracy.

We originally identified twelve study designs (randomized controlled trial, basic sciences studies, case series, case-control study, cohort, comparative study, cross-sectional study, medico-economics study, simulation study, mixed methods study, qualitative study, systematic review & meta-analysis) and regrouped them under nine categories (randomized controlled trial, basic sciences studies, case-control study, cohort, comparative study, cross-sectional study, medico-economics study/simulation study, mixed/qualitative study, systematic review & meta-analysis).

**Supplementary material 5.** Strategy for extracting the sample size.

We treated the task of identifying the sample sizes as a question answering (QA) task. We examined various transformer models (e.g., BioBERT-SQuAD, ScieBERT-SQuAD) and finally selected the ELECTRA-large model trained on PubMed abstracts and then on SQuAD2 dataset [1], which performed the best. We fine-tuned this model on a subset of the PICO training dataset [2] for participant information, which required some reformatting (in this PICO corpus, the sentences of MEDLINE abstracts are annotated with Participants/Problem (P), Intervention (I), Comparison (C) and Outcome (O), we used the participants section and reformatted the annotations into questions and answers for training our QA model). We applied the fine-tuned model to the abstracts (the first 10,000 characters) and if the extracted sample size had a low probability (less than .5), the model was then applied to the full text. The case control studies described two sample sizes; therefore, we kept the extracted two sample sizes with the highest probabilities for case control studies as opposed to the other studies where only one most probable sample size was extracted. Finally, all the extracted numeral words were converted to numeric digits using the word2number python package. The QA model was implemented using Hugging Face library [3]. The performance of the model was examined on a held of biomedical journal data (test set prepared by our experts) and achieved an accuracy of 78%. We used the following four categories of sample size 1) ≤100 or missing sample size, 2) 101-1000, 3) 1001-10 000 and 4) >10 000.

**Supplementary Table 3.** Description of cases and controls by editorial, authorship, and study characteristics for the CMI journal (n=320).

| Variables | Cases n (%) | Controls n (%) |
|---|---|---|
| ***Authorship characteristics*** | | |
| First author gender[a] *(Missing)*, n (%) | *2 (1.2)* | *13 (8.1)* |
| Woman | 82 (51.3) | 53 (33.1) |
| Man | 76 (47.5) | 94 (58.8) |
| Last author gender[a] *(Missing)*, n (%) | *10 (6.3)* | *15 (9.4)* |
| Woman | 35 (21.9) | 37 (23.1) |
| Man | 115 (71.9) | 108 (67.5) |
| Mean number of authors (±SD, Q2: Q1-Q3) | 9.96 (±5.18, 9.5:7-12) | 8.82 (±4.15,8:6-11) |
| Mean percentage of women authors[a] (±SD, Q2: Q1-Q3) | 39.7 (±22.9, 40.0:21.1-55.6) | 38.7 (±23.8, 37.5:22.2-57.1) |
| Proportion of women co-authors on byline[a] | | |
| 0% women | 13 (8.1) | 17 (10.6) |
| Up to 49% women | 83 (51.9) | 89 (55.6) |
| From 50 to 99% women | 63 (39.4) | 50 (31.3) |
| 100% women | 1 (0.6) | 4 (2.5) |
| ***Editorial process variables*** | | |
| Editor gender[a], n (%) | | |
| Woman | 28 (22.1) | 38 (28.6) |
| Man | 99 (77.9) | 95 (71.4) |
| Mean delay to last decision (±SD, Q2: Q1-Q3), days | 103.5 (±42.9, 100:71-133) | 39.2 (±30.6, 34:22-51) |
| ***Study characteristics*** | | |
| Study design, n (%) | | |
| Randomized controlled trial | 10 (6.9) | 1 (0.7) |
| Basic sciences studies | 10 (6.9) | 11 (7.3) |
| Case-control study | 14 (9.6) | 9 (5.9) |
| Cohort study | 37 (25.3) | 38 (25.2) |
| Comparative study | 42 (28.8) | 61 (40.4) |
| Cross-sectional study | 19 (13.0) | 19 (12.6) |
| Medico-economics/simulation study | 5 (3.4) | 2 (1.3) |
| Mixed/qualitative methods | 3 (2.0) | 1 (0.7) |
| Systematic review & meta-analysis | 6 (4.1) | 9 (5.9) |
| Sample size, n (%) | | |
| No sample size | 4 (2.5) | 2 (1.3) |
| ≤100 | 40 (25.0) | 48 (30.0) |
| 101-1000 | 90 (56.3) | 76 (47.5) |
| 1001-10000 | 18 (11.3) | 24 (15.0) |
| >10000 | 8 (5.0) | 10 (6.3) |
| Type of funding, n (%) | | |
| No funding | 9 (5.6) | 6 (3.7) |
| No information of the funding | 70 (43.8) | 80 (50.0) |
| Non-profit | 28 (17.5) | 29 (18.1) |
| Associations and foundations | 12 (7.5) | 12 (7.5) |
| For profit | 2 (1.2) | 2 (1.2) |
| International organizations | 4 (2.5) | 4 (2.5) |
| Non-profit & associations and foundations | 23 (14.4) | 21 (13.1) |
| Non-profit & for-profit | 12 (7.5) | 6 (3.7) |

[a]Gender identified with an accuracy above 60%

**Supplementary Table 4.** Acceptance for publication in a random sample of 7,000 accepted/rejected manuscripts submitted to 21 BMJ Publishing Group journals between January 1, 2012 and December 31, 2019 with at least two coauthors on the byline . Multivariable analyses from three sensitivity analyses.

| Independent variables | Multivariable analysis[a,b,c] | | | Multivariable analysis[d,e,f] | | | Multivariable analysis[g,h,i] | | |
|---|---|---|---|---|---|---|---|---|---|
| | OR | 95%CI | p-value | OR | 95%CI | p-value | OR | 95%CI | p-value |
| **Author / Editor characteristics** | | | | | | | | | |
| First author's gender[c] (ref= Man) | | | 0.645 | | | 0.718 | | | 0.309 |
| Woman | 1.03 | (0.92 to 1.14) | | 1.02 | (0.91 to 1.15) | | 1.06 | (0.95 to 1.17) | |
| Last author's gender[c] (ref= Man) | | | 0.454 | | | 0.338 | | | 0.569 |
| Woman | 0.96 | (0.86 to 1.07) | | 0.94 | (0.84 to 1.06) | | 0.97 | (0.87 to 1.08) | |
| First author's geographical affiliation[e] (ref=Europe) | | | <0.001 | | | <0.001 | | | <0.001 |
| Africa | 0.87 | (0.51 to 1.49) | 0.616 | 0.70 | (0.39 to 1.26) | 0.126 | 0.87 | (0.53 to 1.43) | 0.586 |
| Asia | 0.56 | (0.46 to 0.69) | <0.001 | 0.56 | (0.45 to 0.71) | <0.001 | 0.61 | (0.51 to 0.73) | <0.001 |
| North America | 1.09 | (0.96 to 1.25) | 0.198 | 1.06 | (0.92 to 1.22) | 0.441 | 1.08 | (0.96 to 1.23) | 0.206 |
| Oceania | 1.09 | (0.88 to 1.35) | 0.429 | 1.13 | (0.90 to 1.43) | 0.280 | 1.05 | (0.85 to 1.28) | 0.666 |
| South America | 0.77 | (0.48 to 1.24) | 0.284 | 0.68 | (0.40 to 1.15) | 0.147 | 0.78 | (0.51 to 1.21) | 0.271 |
| First author's country of affiliation income (ref=high income) | | | 0.003 | | | 0.058 | | | <0.001 |
| Upper middle income | 0.60 | (0.45 to 0.81) | 0.001 | 0.68 | (0.48 to 0.96) | 0.027 | 0.60 | (0.48 to 0.77) | <0.001 |
| Lower middle and low income | 0.73 | (0.50 to 1.06) | 0.096 | 0.71 | (0.48 to 1.07) | 0.099 | 0.69 | (0.49 to 0.97) | 0.032 |
| Geographical/income diversity between first/last authors (ref=same country) | | | <0.001 | | | <0.001 | | | <0.001 |
| Different countries but same geographical/income groups | 1.37 | (1.07 to 1.77) | 0.014 | 1.34 | (1.03 to 1.95) | 0.029 | 1.35 | (1.06 to 1.71) | 0.015 |
| Different countries and geographical affiliation but same income group | 1.44 | (1.12 to 1.84) | 0.004 | 1.50 | (1.16 to 1.95) | 0.002 | 1.37 | (1.09 to 1.72) | 0.007 |
| Different income groups | 1.55 | (1.14 to 2.09) | 0.005 | 1.72 | (1.24 to 2.39) | 0.001 | 1.53 | (1.17 to 2.01) | 0.002 |
| **Study characteristics** | | | | | | | | | |
| Study design (ref= Randomized controlled trial) | | | 0.091 | | | 0.056 | | | 0.0185 |
| Basic sciences studies | 1.49 | (1.04 to 2.15) | 0.031 | 1.55 | (1.04 to 2.30) | 0.030 | 1.40 | (1.01 to 1.94) | 0.046 |
| Case-control study | 0.92 | (0.62 to 1.39) | 0.705 | 0.89 | (0.59 to 1.36) | 0.596 | 0.88 | (0.61 to 1.29) | 0.522 |
| Cohort study | 0.87 | (0.69 to 1.09) | 0.221 | 0.84 | (0.66 to 1.06) | 0.139 | 0.81 | (0.66 to 1.01) | 0.060 |
| Comparative study | 0.89 | (0.68 to 1.18) | 0.422 | 0.84 | (0.63 to 1.13) | 0.251 | 0.89 | (0.68 to 1.15) | 0.354 |
| Cross-sectional study | 0.87 | (0.67 to 1.12) | 0.285 | 0.83 | (0.64 to 1.09) | 0.187 | 0.82 | (0.65 to 1.05) | 0.111 |
| Medico-economics/simulation study | 0.91 | (0.52 to 1.60) | 0.740 | 0.81 | (0.45 to 1.46) | 0.486 | 0.87 | (0.52 to 1.48) | 0.610 |
| Mixed/qualitative methods | 0.88 | (0.68 to 1.14) | 0.339 | 0.84 | (0.64 to 1.10) | 0.196 | 0.83 | (0.66 to 1.06) | 0.137 |
| Systematic review & meta-analysis | 0.82 | (0.58 to 1.17) | 0.275 | 0.81 | (0.56 to 1.16) | 0.246 | 0.79 | (0.57 to 1.10) | 0.165 |

| | | | | | | | | | |
|---|---|---|---|---|---|---|---|---|---|
| Sample size (ref= ≤100 or no sample size) | | | 0.222 | | | 0.072 | | | 0.119 |
| 101-1000 | 1.06 | (0.94 to 1.21) | 0.355 | 1.11 | (0.97 to 1.27) | 0.118 | 1.07 | (0.95 to 1.20) | 0.262 |
| 1001-10000 | 1.08 | (0.92 to 1.27) | 0.334 | 1.07 | (0.90 to 1.27) | 0.438 | 1.10 | (0.95 to 1.28) | 0.210 |
| >10000 | 1.22 | (1.01 to 1.48) | 0.038 | 1.29 | (1.06 to 1.58) | 0.012 | 1.24 | (1.04 to 1.47) | 0.018 |
| Type of funding (ref. No funding) | | | <0.001 | | | <0.001 | | | <0.001 |
| No information of funding | 1.10 | (0.91 to 1.33) | 0.342 | 1.06 | (0.86 to 1.29) | 0.584 | 1.06 | (0.89 to 1.27) | 0.494 |
| Non-profit | 1.17 | (0.95 to 1.45) | 0.139 | 1.14 | (0.91 to 1.42) | 0.268 | 1.18 | (0.97 to 1.43) | 0.104 |
| Associations and foundations | 1.49 | (1.13 to 1.96) | 0.004 | 1.37 | (1.03 to 1.84) | 0.031 | 1.45 | (1.12 to 1.87) | 0.004 |
| For-profit | 1.15 | (0.81 to 1.63) | 0.427 | 1.16 | (0.81 to 1.67) | 0.425 | 1.11 | (0.80 to 1.54) | 0.519 |
| International organizations | 2.48 | (1.74 to 3.55) | <0.001 | 2.67 | (1.82 to 3.92) | <0.001 | 2.26 | (1.63 to 3.14) | <0.001 |
| Non-profit & associations & foundations | 1.28 | (1.05 to 1.58) | 0.016 | 1.24 | (1.00 to 1.54) | 0.050 | 1.30 | (1.07 to 1.57) | 0.007 |
| Non-profit & for-profit | 1.20 | (0.86 to 1.67) | 0.296 | 1.18 | (0.83 to 1.69) | 0.362 | 1.31 | (0.97 to 1.78) | 0.083 |

[a]Model performed for manuscripts with more than one author (n=5,873 observations). [b]Model adjusted on the speciality of the research topic (p=0.774). [c]Gender determined with accuracy ≥70%. [d]Model performed for manuscripts with more than one author (n=5,220 observations). [e]Model adjusted on the speciality of the research topic (p=0.823). [f]Gender determined with accuracy ≥80%. [g]Model performed for manuscripts with more than one author (n=6,828 observations). [h]Model adjusted on the speciality of the research topic (p=0.653). [i]Gender determined by multiple imputation.
